# Impact of SARS-CoV-2 antibodies at delivery in women, partners and newborns

**DOI:** 10.1101/2020.09.14.20191106

**Authors:** Pia Egerup, Line Fich Olsen, Ann-Marie Hellerung Christiansen, David Westergaard, Elin Rosenbek Severinsen, Kathrine Vauvert Römmelmayer Hviid, Astrid Marie Kolte, Amalie Dyhrberg Boje, Marie-Louise Mathilde Friis Bertelsen, Lisbeth Prætorius, Anne Zedeler, Josefine Reinhardt Nielsen, Didi Bang, Sine Berntsen, Jeppe Ethelberg-Findsen, Ditte Marie Storm, Judith Bello-Rodríguez, Andreas Ingham, Joaquim Ollé-López, Eva R Hoffmann, Charlotte Wilken-Jensen, Lone Krebs, Finn Stener Jørgensen, Henrik Westh, Henrik Løvendahl Jørgensen, Nina la Cour Freiesleben, Henriette Svarre Nielsen

## Abstract

**Background:** Only few studies have focused on serological testing for SARS-CoV-2 in pregnant women and no previous study has investigated the frequency in partners. The aim was to investigate the frequency and impact of SARS-CoV-2 in parturient women, their partners and newborns.

**Methods:** From April 4^th^ to July 3^rd^, 2020, all parturient women, their partners and newborns were invited to participate in the study. Participating women and partners had a pharyngeal swab and a blood sample taken at admission and immediately after delivery a blood sample was drawn from the umbilical cord. The swabs were analyzed for SARS-CoV-2 RNA by PCR and the blood samples were analyzed for SARS-CoV-2 antibodies. Full medical history, obstetric- and neonatal information were available.

**Results:** A total of 1,361 parturient women, 1,236 partners and 1,342 newborns participated in the study. No associations between previous COVID-19 disease and obstetric- or neonatal complications were found. The adjusted serological prevalence was 2.9% in women and 3.8% in partners. The frequency of blood type A was significantly higher in women with antibodies compared to women without antibodies. 17 newborns had SARS-CoV-2 IgG antibodies, and none had IgM antibodies. Full serological data from 1,052 families showed an absolute risk of infection of 0.37 if the partner had antibodies. Only 55% of individuals with antibodies reported symptoms.

**Conclusion:** This large prospective cohort study reports no association between COVID-19 and obstetric- or neonatal complications. The family pattern showed a substantial increase in absolute risk for women living with a partner with antibodies.

## Introduction

Coronavirus disease 2019 (COVID-19) caused by severe acute respiratory syndrome coronavirus 2 (SARS-CoV-2) is a worldwide pandemic mainly transmitted by droplets. However, other transmission routes have been postulated, including vertical transmission from mother to fetus.^1^ Currently, data from studies investigating the impact of COVID-19 in pregnancy have mainly been based on detection of viral RNA by polymerase chain reaction (PCR) analysis.^2,3^ A recent study of 675 women admitted for delivery in New York City found a prevalence of COVID-19 of 10.4%.^3^ Of these 78.6% were asymptomatic.^3^ The authors found a significantly higher incidence of cesarean sections, postpartum complications and placental pathology among women with COVID-19 compared to women without the disease.^3^

To date only a few studies have focused on serological testing for SARS-CoV-2 in pregnant women.^4-6^ A recent study from Philadelphia, USA, performed serological testing on 1,293 women admitted for delivery and found a seroprevalence of 6.2%.^6^ The study, however, did not include comprehensive data concerning obstetric and neonatal outcomes. Two smaller case reports from China documented SARS-CoV-2 antibodies in newborns with COVID-19 positive mothers indicating possible vertical transmission.^4,7^ To our knowledge, no previous study has investigated the frequency of SARS-CoV-2 in partners of pregnant women.

This study aimed to investigate the frequency and impact of SARS-CoV-2 in parturient women, their partners and newborns.

We here report the results of a prospective cohort study with unselected serological testing in 1,313 parturient women, 1,189 partners and 1,206 newborns to identify if SARS-CoV-2 infection is associated with obstetric and neonatal complications. The study included 1,052 families with full serological data.

## Methods

### Participants

The Department of Obstetrics and Gynecology at Copenhagen University Hospital Hvidovre serves a broad population and has 7000 births per year. From April 4^th^, 2020 to July 3^rd^, 2020, (3 months), all women giving birth at the department, their partners and newborns were invited to participate in the study.

### Sample collection

Participating women and their partners had a pharyngeal swab and a blood sample taken at admission. Immediately after delivery, an umbilical cord blood sample was drawn if the newborn participated in the study.

### Sample analysis

The pharyngeal swabs from the participating women and their partners were analyzed for virus SARS-CoV-2 RNA by PCR testing. The serum from the blood samples from women, partners and newborns were analyzed for SARS-CoV-2 antibodies (IgM and IgG).

Samples were analyzed using YHLO’s iFlash 1800 and SARS-CoV-2 IgM/IgG kits. Following a recent study on the specific assay, a negative result was defined as IgM < 8 AU/mL and IgG < 10 AU/ml and a positive result was defined as values >= 8 AU/mL for IgM and >= 10 AU/mL for IgG, respectively. This has been demonstrated to have a sensitivity of 42.0% and a specificity of 99.7% for IgM and 94.0% and 99.3% for IgG, respectively.^8^

### Pregnancy outcomes

Baseline characteristics of the included women and the obstetric outcomes were recorded by cross-referencing with the electronic health records. Baseline characteristics included age, pre-pregnancy body mass index (BMI), smoking, alcohol, chronic diseases, fertility treatment, parity and information on previous pregnancies. Outcomes related to pregnancy included pregnancy complications (e.g. gestational diabetes (GDM), gestational hypertension, preeclampsia, eclampsia, premature rupture of membranes (PPROM), group B streptococcus in urine (GBS)) and obstetric outcomes (e.g. gestational week, preterm birth, vaginal delivery, vacuum extraction, cesarean section, induced labor, postpartum bleeding, fever, antibiotic treatment during birth, duration of hospitalization). Additionally, a short questionnaire concerning symptoms of COVID-19 since December 2019 and lung disease was completed by all participating women.

## Included partners

Characteristics of the included partners were obtained through a short questionnaire and by cross-referencing with the electronic health records. The recorded information included age, BMI, smoking, chronic diseases, symptoms of COVID-19, previously diagnosed COVID-19, and travels outside Denmark since December 2019.

### Neonatal outcomes

The neonatal outcomes included birth information (gestational week at birth, meconium stained amniotic fluid), sex, birth weight, birth length, head and abdominal circumference, Apgar scores, umbilical arterial pH, base excess, malformations, CPAP need and admission to the neonatal department. The information was recorded by cross-referencing with the newborn’s electronic health records.

### Statistical analysis

Data were analyzed using the statistical software R. Normally distributed comparison between groups was performed using a t-test, and for non-normally distributed data the Wilcox rank-sum test was used. Categorical data was analyzed using the Fisher’s exact test. Calculation of adjusted prevalence, odds ratios, attributable risk, and confidence intervals was performed using the R package epiR.^9^ Observed prevalence was adjusted for the known specificity and sensitivity of the assays^8^, and confidence intervals were established using the Wilson method.

Multiple regression was performed using the R package rms.^10^ A p-value < 0.05 was considered statistically significant. DW performed the statistical analysis.

### Ethical approval

The study was approved by the Knowledge Centre for Data Protection and Compliance, The Capital Region of Denmark (P-2020-255) and by the Scientific Ethics Committee of the Capital Region of Denmark (journal number H-20022647). All participants provided written informed consent. Informed consent for the newborn was obtained via informed consent from both parents or from the mother alone in case of sole parental custody.

## Results

A total of 1,810 women gave birth at the hospital in the study period, of which 1,361 (75.2%) participated in the study (Figure 1). Furthermore, 1,236 partners and 1,342 newborns participated.

**Figure 1.**
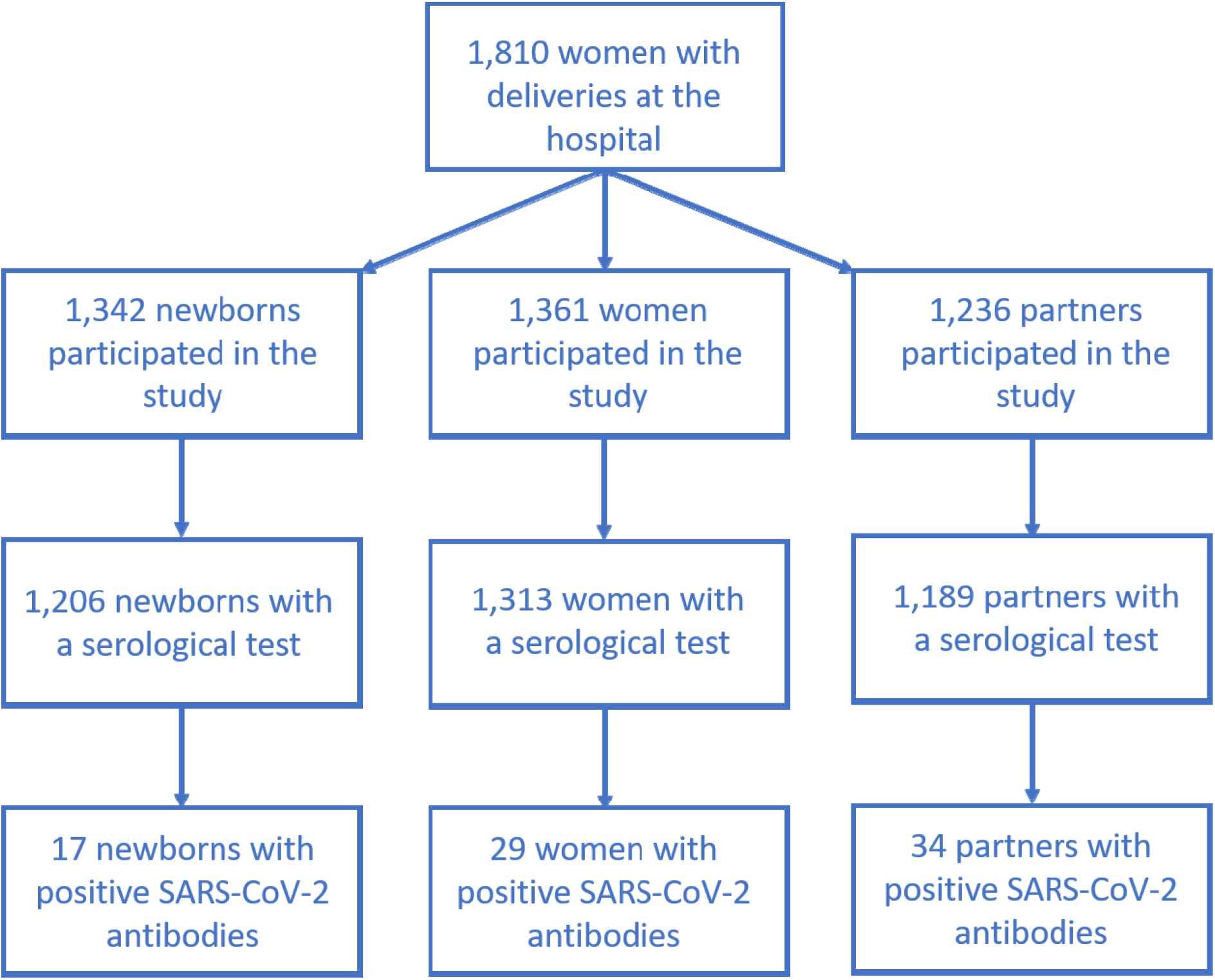
Flowchart illustrating the study population.

Of the included parturient women, 29 had antibodies against SARS-CoV-2 (2.2%) (Figure 1). The adjusted prevalence was 2.85% (95%CI 1.87%-4.25%), when taking into account the assay specificity and sensitivity. Table 1 shows the characteristics of the included parturient women according to their SARS-CoV-2 antibody status. The median age was 31.5 (IQR 28.6-34.7) years in the negative group and 31.4 (IQR 28.0-33.3) years in the positive group with no significant difference (p=0.3). There was no significant difference between pre-pregnancy characteristics in relation to SARS-CoV-2 antibodies, except blood type and that women with antibodies reported more COVID-19-like symptoms (p=0.025). Nonetheless, only 52% (95%CI 33%-71%) of women with antibodies reported symptoms. Significantly more women with SARS-CoV-2 antibodies had blood type A compared to the negative group (p=0.025). Only one woman had a positive pharyngeal swab. Four women in the positive group had previously been diagnosed with COVID-19. One woman with negative SARS-CoV-2 antibodies had a positive COVID-19 swab 4 days before delivery. The median gestational age at birth was 40+1 weeks (281 days) for both groups. There was no significant difference in outcomes or obstetric complications between the two groups. A total of 14 newborns (64%) had positive SARS-CoV-2 in the positive group, and three newborns (0.3%) in the negative group. There was a significant increase in the relative risk for newborns to have antibodies if the mother had antibodies (RR=446, 95%CI 110-1807, p<0.001). This corresponded to an 86% increase in absolute risk (95%CI 69%-104%, p < 0.001).

**Table 1.**
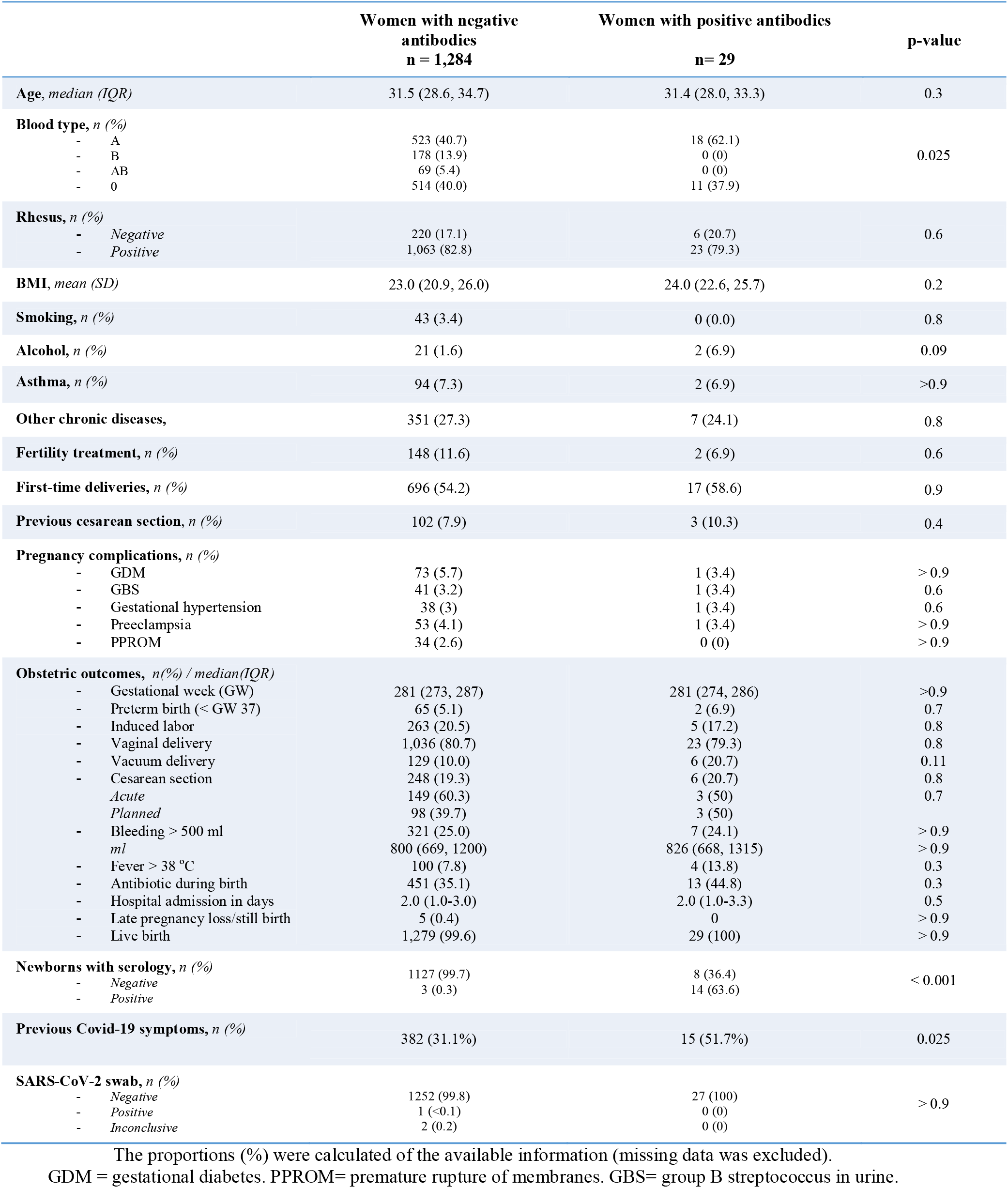
Characteristics of the included parturient women according to SARS-CoV-2 antibodies

Cohort characteristics of the included partners according to SARS-CoV-2 antibodies are presented in Table 2. A total of 34 partners had antibodies against SARS-CoV-2 (2.9%). The adjusted prevalence was 3.8% (95%CI 2.6%-5.5%). There was no significant difference in baseline characteristics (e.g. age, BMI, smoking, lung disease) except chronic diseases (p=0.045). There was no increased history of travels outside Denmark since December 2019 in the positive group. Significantly more men in the positive group had a previous positive SARS-CoV-2 swab (p<0.001) and reported more symptoms (p<0.001). However, only 61% (95%CI 42%-77%) of partners with SARS-CoV-2 antibodies reported any symptoms.

**Table 2.**
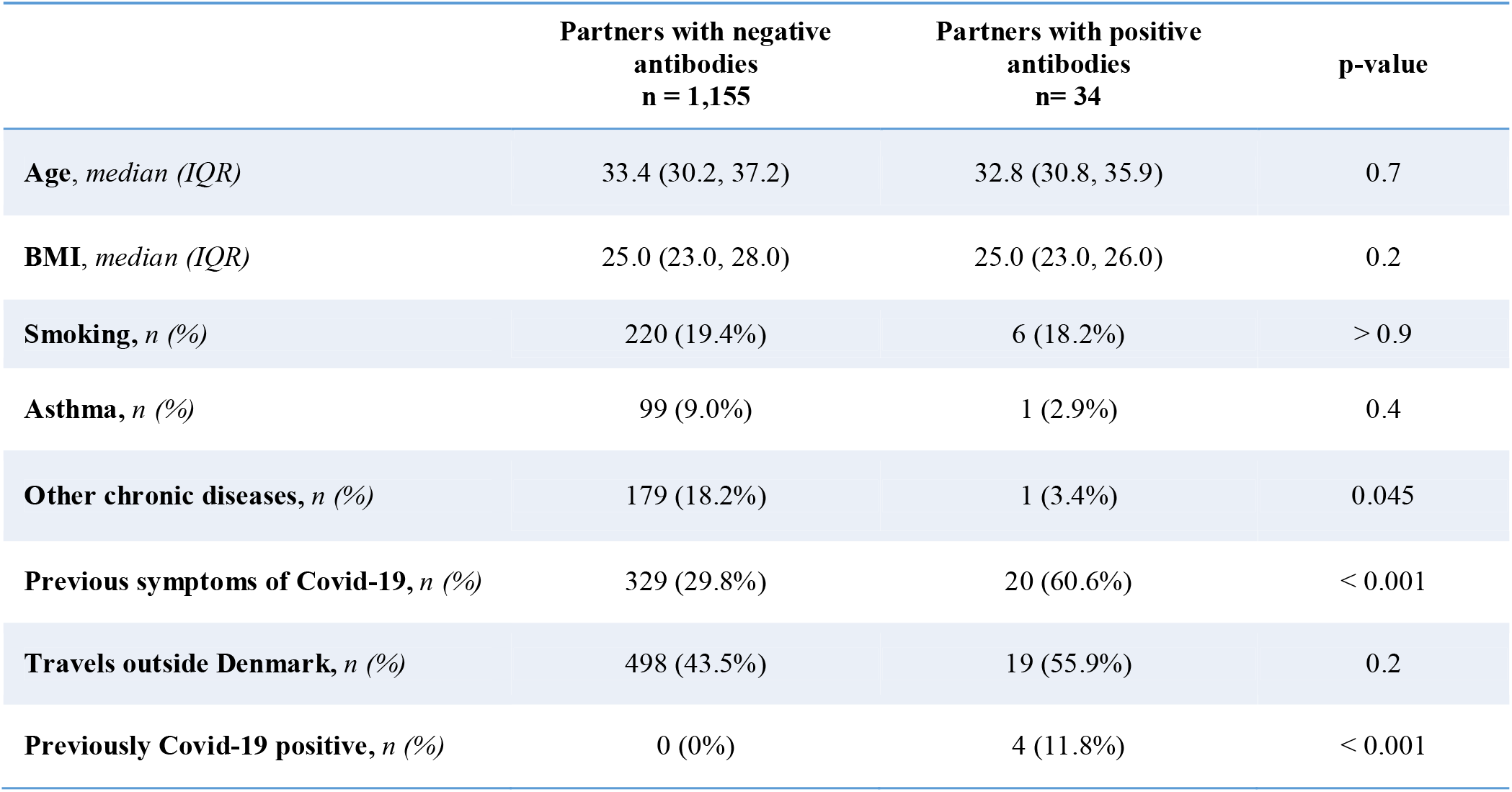
Characteristics of the included partners according to SARS-CoV-2 antibodies

|  | Partners with negative<br>antibodies<br>n = 1,155 | Partners with positive<br>antibodies<br>n= 34 | p-value |
| --- | --- | --- | --- |
| **Age, median (IQR)** | 33.4 (30.2, 37.2) | 32.8 (30.8, 35.9) | 0.7 |
| **BMI, median (IQR)** | 25.0 (23.0, 28.0) | 25.0 (23.0, 26.0) | 0.2 |
| **Smoking, n (%)** | 220 (19.4%) | 6 (18.2%) | > 0.9 |
| **Asthma, n (%)** | 99 (9.0%) | 1 (2.9%) | 0.4 |
| **Other chronic diseases, n (%)** | 179 (18.2%) | 1 (3.4%) | 0.045 |
| **Previous symptoms of Covid-19, n (%)** | 329 (29.8%) | 20 (60.6%) | < 0.001 |
| **Travels outside Denmark, n (%)** | 498 (43.5%) | 19 (55.9%) | 0.2 |
| **Previously Covid-19 positive, n (%)** | 0 (0%) | 4 (11.8%) | < 0.001 |

None of the women or partners with antibodies in our cohort had been hospitalized for COVID-19.

Table 3 shows the characteristics of the newborns. Of the 1,206 newborns with serological test, 17 had positive antibodies against SARS-CoV-2 (1.4%). All were IgG positive and IgM negative. A total of 5.0% and 5.9% of the newborns were born prematurely in the negative and positive group, respectively (p=0.6). There was no significant difference according to meconium-stained amniotic fluid, sex, birth length, head circumference, abdominal circumference, umbilical arterial pH, or Apgar scores. The frequency of neonatal complications (malformations, CPAP need or admission to a neonatal department) was not significantly different between the two groups. The median birth weight in the negative newborns was 3,528 g (IQR 3,200-3,860) compared to 3,800 g (IQR 3,550-4,106) in positive newborns. Birth weight was significantly different between the two groups (p=0.015) but this association was rendered non-significant when corrected for gestational age and newborn birth length (p=0.18). Except for two SARS-CoV-2 negative newborns all were alive at discharge.

**Table 3.** Characteristics of the included newborns according to SARS-CoV-2 antibodies

|  | Newborns with negative<br>antibodies<br>n = 1,189 | Newborns with positive<br>antibodies<br>n= 17 | p-value |
| --- | --- | --- | --- |
| **Gestational week at birth, median (IQR)** | 281 (273, 287) | 284 (278, 289) | 0.14 |
| **Premature babies (gestational week < 37), n (%)** | 60 (5.0) | 1 (5.9) | 0.6 |
| **Meconium-stained amniotic fluid, n (%)** | 190 (16.1) | 2 (11.8) | > 0.9 |
| **Sex, n (%)** |  |  |  |
| - Female | 623 (52.4) | 10 (58.8) | 0.6 |
| - Male | 566 (47.6) | 7 (41.2) |  |
| **Birth weight in g, median (IQR)** | 3,528 (3,200, 3,860) | 3,800 (3,550, 4,106) | 0.015 |
| **Low birth weight babies < 2500g, n (%)** | 47 (4.0) | 0 (0) | >0.9 |
| **Length in cm, median (IQR)** | 52.00 (50.00, 53.00) | 53.00 (51.00, 53.00) | 0.12 |
| **Head circumference in cm, median (IQR)** | 35.00 (34.00, 36.00) | 35.00 (34.00, 37.00) | 0.1 |
| **Abdominal circumference in cm, median (IQR)** | 33.00 (32.00, 34.00) | 33.00 (33.00, 35.00) | 0.3 |
| **APGAR-1, median (IQR)** | 10.00 (9.00, 10.00) | 10.00 (9.00, 10.00) | 0.3 |
| **APGAR-5, median (IQR)** | 10.00 (10.00, 10.00) | 10.00 (10.00, 10.00) | > 0.9 |
| **APGAR-5 < 7, n (%)** | 4 (0.3) | 0 (0.0) | > 0.9 |
| **Arterial pH, median (IQR)** | 7.23 (7.17, 7.29) | 7.23 (7.20, 7.24) | 0.7 |
| **Arterial pH < 7.0, n (%)** | 9 (0.9) | 0 (0.0) | > 0.9 |
| **Base excess, median (IQR)** | -4.5 (-6.8, -2.3) | -6.3 (-7.9, -4.5) | 0.051 |
| **CPAP, n (%)** | 160 (13.5) | 2 (11.8) | > 0.9 |
| **Malformation, n (%)** | 21 (1.8) | 1 (5.9) | 0.3 |
| **Hospital admission in days, median (IQR)** | 2.00 (1.00, 3.00) | 1.00 (1.00, 4.00) | 0.9 |
| **Neonatal admission, n (%)** | 132 (11.1) | 2 (11.8) | > 0.9 |
| **Alive at discharge, n (%)** |  |  |  |
| - Yes | 1175 (99.8) | 17 (100) | >0.9 |
| - No | 2 (0.2) | 0 (0) |  |
| **Mothers with antibodies, n (%)** |  |  |  |
| - Negative | 1142 (99.4) | 3 (17.6) | <0.001 |
| - Positive | 7 (0.6) | 14 (82.4) |  |

Looking at family patterns of infection across 1,052 families with complete serological data, we found seven families where all members had antibodies against SARS-CoV-2 (Supplemental Table 1). The patterns of familial infection can be seen in Figure 2. We found that there was a 45.7% (95%CI 23.2%-68.2%, p<0.001) increase in the absolute risk of antibody positivity for mothers living with a partner that had SARS-CoV-2 antibodies. The absolute risk of infection was 0.37 (95%CI 0.19-0.55) for the woman if her partner tested positive.

**Figure 2.**
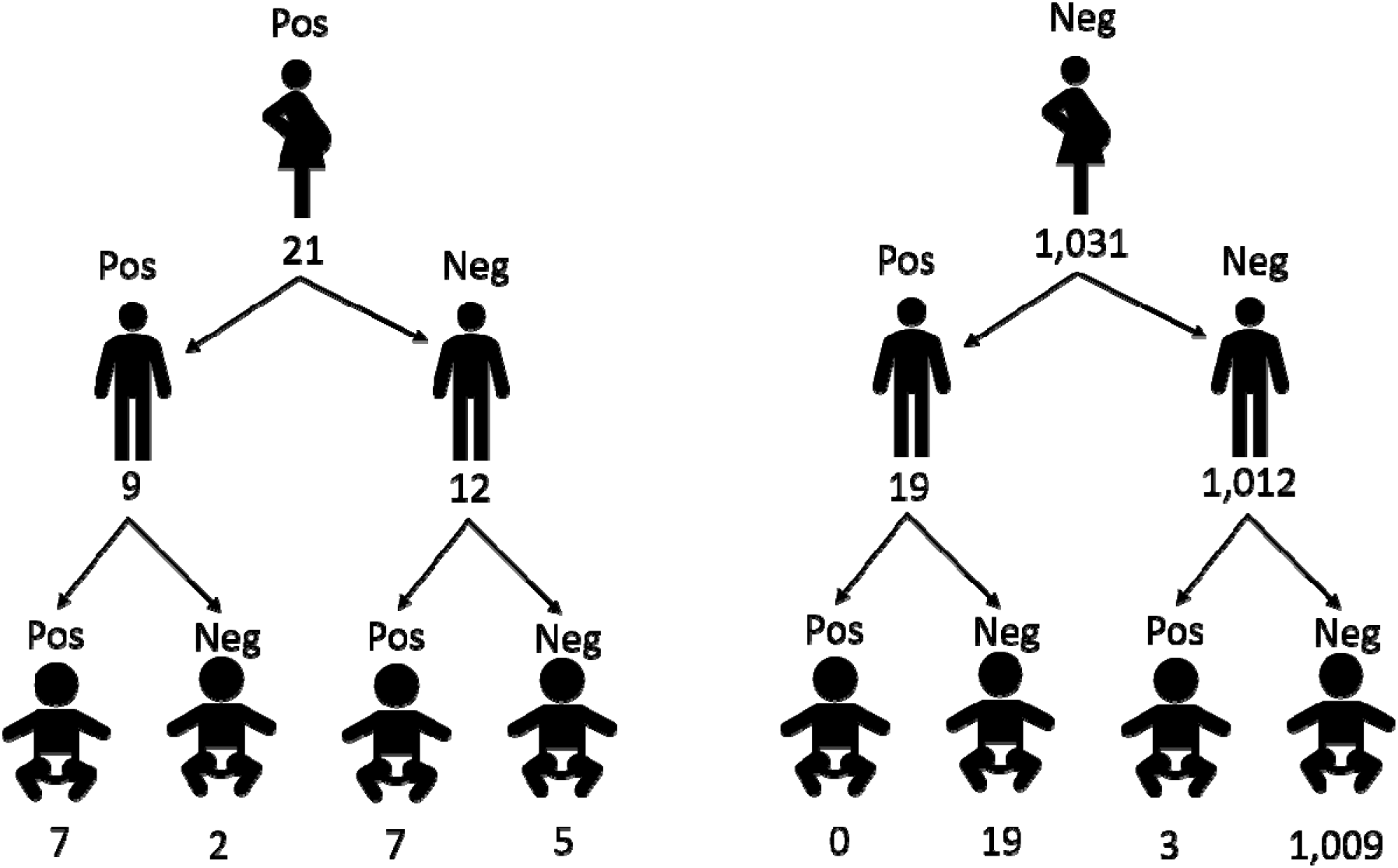
Family pattern of SARS-CoV-2 transmission

## Discussion

In this prospective cohort study including 1,361 parturient women, 1,236 partners and 1,342 newborns, we found no association between previous COVID-19 disease and obstetric- or neonatal complications. The adjusted serological prevalence was 2.9% in parturient women and 3.8% in partners. None of the antibody-positive women or partners in our cohort had previously been hospitalized for COVID-19. No newborns had positive IgM antibodies against SARS-CoV-2. Full serological data from 1,052 families showed an absolute risk of infection of 0.37 for the women if the partner was positive. Only 55% of individuals with antibodies reported any symptoms of COVID-19 disease (no difference between women and partners).

A recently published study of 675 parturient women from New York City found a significantly higher frequency of cesarean delivery of 46.7% in symptomatic COVID-19 women as opposed to 45.5% in asymptomatic COVID-19 women and 30.9% in women without COVID-19.^3^ In our cohort, approximately 20% of the included women had a cesarean delivery, similar to women with and without SARS-CoV-2 antibodies. In the study by Prabhu et al., however, they used nasopharyngeal swab and RT-PCR analysis to determine COVID-19 infection.^3^ Thus, this reflected acute COVID-19 disease. The authors state, however, that the obstetric management was not altered according to symptoms or COVID-19 status, and the indication for cesarean delivery was not statistically different according to COVID-19 status (p=0.83).^3^ The difference in frequency of cesarean delivery between our study and the study by Prabhu et al. may be explained by difference in baseline characteristic of the included women (e.g. less women in our cohort had BMI > 30) or general national differences in obstetric management. Two additional studies have reported a higher frequency of cesarean delivery of 59% and 93%, respectively, in symptomatic COVID-19 positive women.^2,11^ However, the women in these studies were symptomatic and can therefore not be directly compared to our cohort.

The significant difference in blood type with a higher frequency of blood type A in the positive women compared to the negative women has been documented previously.^12^

Our study identified 17 newborns with antibodies against SARS-CoV-2. However, none of the newborns had IgM antibodies. From the beginning of the second trimester, IgG is passively transferred across the placenta from the mother to the fetus. In contrast, IgM antibodies usually are not transferred across the placenta.^13^ The presence of IgG antibodies but not IgM antibodies in the newborns in our study may thus be a result of maternal transfer of antibodies via the placenta and not a vertical transmission of the disease. The SARS-CoV-2 antibody positive newborns with negative mothers may be explained by concentrating of the mother’s antibodies in the newborn. Although infant infection cannot be ruled out, the neonatal outcomes from our study are reassuring. We found no differences in umbilical arterial pH, base excess, CPAP need, Apgar scores, malformations or neonatal hospital admission between IgG positive newborns compared to IgG negative newborns.

Women giving birth are mainly healthy individuals, only hospitalized in relation to the delivery and therefore up to a point represent age-appropriate women in the community. Similarly, testing their partners represents an excellent opportunity to evaluate the prevalence of SARS-CoV-2 in the community. The serological prevalence of people with SARS-CoV-2 antibodies in the Danish population was per May 23^rd^, 2020 estimated to be 1.1% (95%CI 0.5—1.8).^14^ However, only around 40% of the random population sample had been recruited.^14^ The frequency in our cohort was higher, 2.85% (95% CI 1.87%-4.25%) for women and 3.8% (95%CI 2.6%-5.5%) for partners. This may reflect the time difference between the two estimates with more individuals being affected at a later timepoint. The low prevalence of COVID-19 in our cohort compared to other countries^3,5,6^ may be due to the Danish government’s introduction of lockdown measures very early in the epidemic. Serological testing is critical to identify how many individuals have been infected previously and large-scale unselected serological testing should be performed in future studies to evaluate the true prevalence in the community. It is, however, possible that mild cases with affection concentrated to the upper respiratory tract don’t produce antibodies.

A substantial increase in the absolute risk of infection for women living with a partner who had antibodies was documented in our study. A study from China investigating secondary transmission of SARS-CoV-2 in households found that secondary transmission occurred in 41 out of 124 families.^15^ This study, however, included only symptomatic cases. The present study represents asymptomatic or mild cases of COVID-19 and may represent the risk of infection for asymptomatic cases. Absolute risk of infection of 0.37 was found if the partner was positive, i.e. a risk of 37%. However, this risk assumes the parturient women live with their partner, and the primary case in the household is the partner. The pattern of disease in families is highly essential in order to evaluate the risk of infection and to provide guidance to the health authorities in relation to recommendations concerning social distancing. However, as 55% of seropositive participants in our study reported corona-related symptoms, this would require sequential testing among family members with short intervals.

### Strengths and limitations

Our study has several strengths. First, the present study is a comprehensive prospective cohort study from the largest obstetric department in Denmark including both parturient women, partners and newborns. Additionally, the health care system in Denmark is free of charge, which minimizes selection bias, and 95% of all births take place at a hospital. Secondly, we used serological testing, which is critical to investigate the association between previous COVID-19 disease and obstetric- and neonatal complications. Finally, we had complete data on obstetric- and neonatal outcomes from parturient women and their newborns in the study period, and all included women were pregnant when the novel virus spread worldwide, and we can thus conclude that the infection happened during pregnancy.

In general, our study population was young with a median age of 31 years, had a normal BMI, were non-smokers and had had asymptomatic or mild COVID-19 disease without the need for hospitalization. Our results and conclusion may therefore not directly be applied to other populations with a higher BMI, higher frequency of smoking or severe COVID-19 infection.

We used the iFlash 1800 with its IgM/IgG kit for serological testing, which has shown highly accurate results.^16^ A recent study has documented the sensitivity and specificity to be 42.0% and 99.7% for IgM and 94.0% and 99.3% for IgG, respectively.^8^ However, future studies are needed to validate the serological assays especially for use as a screening tool in asymptomatic populations.^17^

## Conclusion

In this prospective cohort study with serological testing of parturient women, partners and newborns we found no association between COVID-19 and obstetric- or neonatal complications. We found a seroprevalence of 2.9% in the parturient women and 3.8% in the partners. The family pattern showed a substantial increase in the absolute risk of infection for women living with a partner who had antibodies.

## Data Availability

Data will not be shared due to personal data.

